# AI-Based Synthetic Data in Biomedicine: A Decade of Growth and a Persistent Translation Gap

**DOI:** 10.64898/2026.09.12.26360876

**Authors:** Navid Asgari, Inãki Fernández Pérez, Gorka Epelde, Linghan Zhang, Lior Horesh, Carl Y. Saab, Mordechai Muszkat, Michal Rosen-Zvi

## Abstract

AI-generated synthetic data are increasingly used to address data scarcity, privacy constraints and experimental limitations in biomedicine, but how far these methods have translated into practice remains unclear. We conducted a systematic mapping and bibliometric analysis of **4,143** publications spanning 2015–2025, combining expert annotation with LLM-assisted classification across data modality, medical domain, paper type, deployment status and research stance. Publication volume grew continuously; **77.8%** of papers were strongly supportive while critical work remained below **1%**. Medical imaging dominated the corpus, consistent with well-characterized transformation-group invariances supporting data augmentation and generative modeling. Highly cited primary research concentrated disproportionately in molecular and pharmaceutical applications, where SE(3)-equivariant architectures and structure-prediction models accelerated generative approaches. Only 27 publications reported operational use; omics and tabular clinical data, lacking well-characterized invariance structures, remained underrepresented. These findings reveal a gap between methodological growth and deployment, motivating investment in evaluation standards, deployment reporting and encoding domain-relevant invariances.

---

Generative artificial intelligence (AI) and machine learning are rapidly reshaping science, industry, and society, enabling advances from content generation to increasingly autonomous decision-making [1–3]. Central to these developments is access to large-scale, high-quality data. However, in healthcare and life sciences (HCLS), data availability is fundamentally constrained by limited sample sizes, fragmentation across institutions, stringent privacy regulations, and the high cost and complexity of data acquisition. These challenges restrict both methodological development and the translation of AI into real-world biomedical applications.

Synthetic data generation has emerged as a critical paradigm for overcoming these limitations, with AI models used to simulate realistic biomedical data across a growing range of settings [4–7]. Beyond addressing data scarcity, it enables augmentation of underrepresented conditions, generation of counterfactual scenarios, and construction of multimodal datasets that would be impractical to collect. More broadly, synthetic data is beginning to reshape biomedical and healthcare processes themselves. In the pharmaceutical industry, *in silico* simulations already complement or partially replace costly experimental procedures and expand the space of candidate molecules, a trend likely to intensify as generative models mature. In clinical settings, digital twins [8–10] and predictive models may reduce reliance on invasive or burdensome diagnostic procedures while enabling simulation of diverse medical scenarios for education and research. Synthetic data is therefore not merely a technical workaround for data-scarce learning, but an emerging foundation for rethinking how healthcare and life science organizations acquire, share, and act on knowledge. Its influence is already extending beyond medicine, as data-driven simulation increasingly shapes domains such as synthetic biology, materials science, and industrial biotechnology.

## Motivation and contributions

Over the past decade, synthetic data generation has emerged as a central response to data scarcity, privacy, and scalability challenges in healthcare and life sciences. Prior work has highlighted the practical and regulatory challenges of deploying synthetic data clinically, including data quality, bias, and trust [11], as well as definitional ambiguities in distinguishing observed, process-driven, and AI-generated data [12]. Existing reviews have mapped key application domains such as simulation, method development, and data sharing [7], and catalogued methods and open-source tools across data modalities [13]. The most direct comparator to the present work is Breugel *et al.* [14], whose 2024 review in *Nature Reviews Bioengineering* synthesized the role of generative AI in producing biomedical data across modalities and provided a qualitative typology of methods spanning generation, application, and trust. We see our contribution as complementary rather than overlapping, in three respects. First, where Breugel *et al*. structure their account around method classes, ours is structured around the empirical landscape of published work, offering what is to our knowledge the first corpus-level quantification of how the field has distributed its effort across modalities, paper types, deployment status, and citation impact across a full decade. Second, we report a volume versus impact asymmetry: medical imaging accounts for 37.8% of the corpus by publication count, while molecular and pharmaceutical applications concentrate the high citation tail among primary research papers. Establishing this requires a corpus large enough to populate a citation distribution for each modality and compare its upper tail against publication volume, which a synthesis organized around exemplar methods does not attempt. Third, only 27 of 4,143 papers in our corpus report a system in operational use. This figure bounds what the literature states rather than what the field has achieved, but that it is only a bound is the finding itself: the peer reviewed record captures the early stages of the research pipeline far better than the late ones, a structural property of the publication system that a review organized around method classes is not positioned to observe.

We situate the modality asymmetry described above within emerging theoretical perspectives on learning from synthetic data. Augmentation, itself a form of synthetic data generation, is most effective when transformations preserve semantic labels through well-defined group invariances such as rotations and translations [15, 16], and recent work characterizes the generalization trade-offs this introduces between invariance, distributional shift, and model dependence [17]. Medical imaging naturally admits such transformation groups, making augmentation both theoretically justified and empirically productive, while omics and tabular data often lack clear invariance structures, limiting the reliability of the same strategies. Molecular data occupies a middle ground here: its SE(3) and permutation symmetries are well-characterized and have been productively embedded in equivariant generative architectures, making it a relative success case compared to omics and tabular clinical data, where the relevant invariance structure remains to be discovered. This asymmetry suggests two productive complements to the standard augmentation playbook. The first is to discover the relevant symmetries rather than assume them, which has become a tractable problem for data with latent group structure [18]. The second is to generalize the algebraic object the model operates over: tensor frameworks built on arbitrary finite groups (the *\*_G_* tensor algebra [19], an extension of the *m*-product family [20]) provide provably optimal equivariant representations and have already been applied to omics [21], where signal is abundant but obvious geometric invariances are scarce. We mention these directions not because they bridge the augmentation gap in non-imaging modalities (as they do not, yet) but because they are concrete examples of the broader move from injecting known invariances to learning or constructing the algebraic structure under which the data is invariant in the first place. A parallel but distinct dynamic has shaped the rise of synthetic molecular data: structure-prediction models such as AlphaFold and its successors in protein design have demonstrated that learning from evolutionary and structural constraints can yield generative models of genuine scientific validity [22–25]. The 2024 Nobel Prize in Chemistry, awarded to Baker, Hassabis, and Jumper for computational protein design and structure prediction, brought exceptional visibility to generative AI as a source of scientifically valid molecular data, attracting a wave of downstream research that is clearly reflected in our corpus. Together, these modality-specific trajectories position synthetic data not merely as a technical workaround, but as an evolving data-centric paradigm, one gaining momentum in biomedicine, yet uneven in its adoption, validation, and integration into real-world practice.

To characterize this landscape, we assembled a corpus of 4,143 PubMed indexed papers published between 2015 and April 2025 and annotated every record along five facets: *paper type*, *medical domain*, *synthetic data type*, *practicality*, and *stance*. Annotation combined multi-round expert labeling by six domain experts with LLM-assisted classification at corpus scale, validated against a held-out set of human annotated papers, and was followed by a bibliometric analysis of citation impact. Figure 1 summarizes the pipeline, from corpus construction through annotation to label consolidation. Full procedures are given in Methods.

**Fig. 1:**
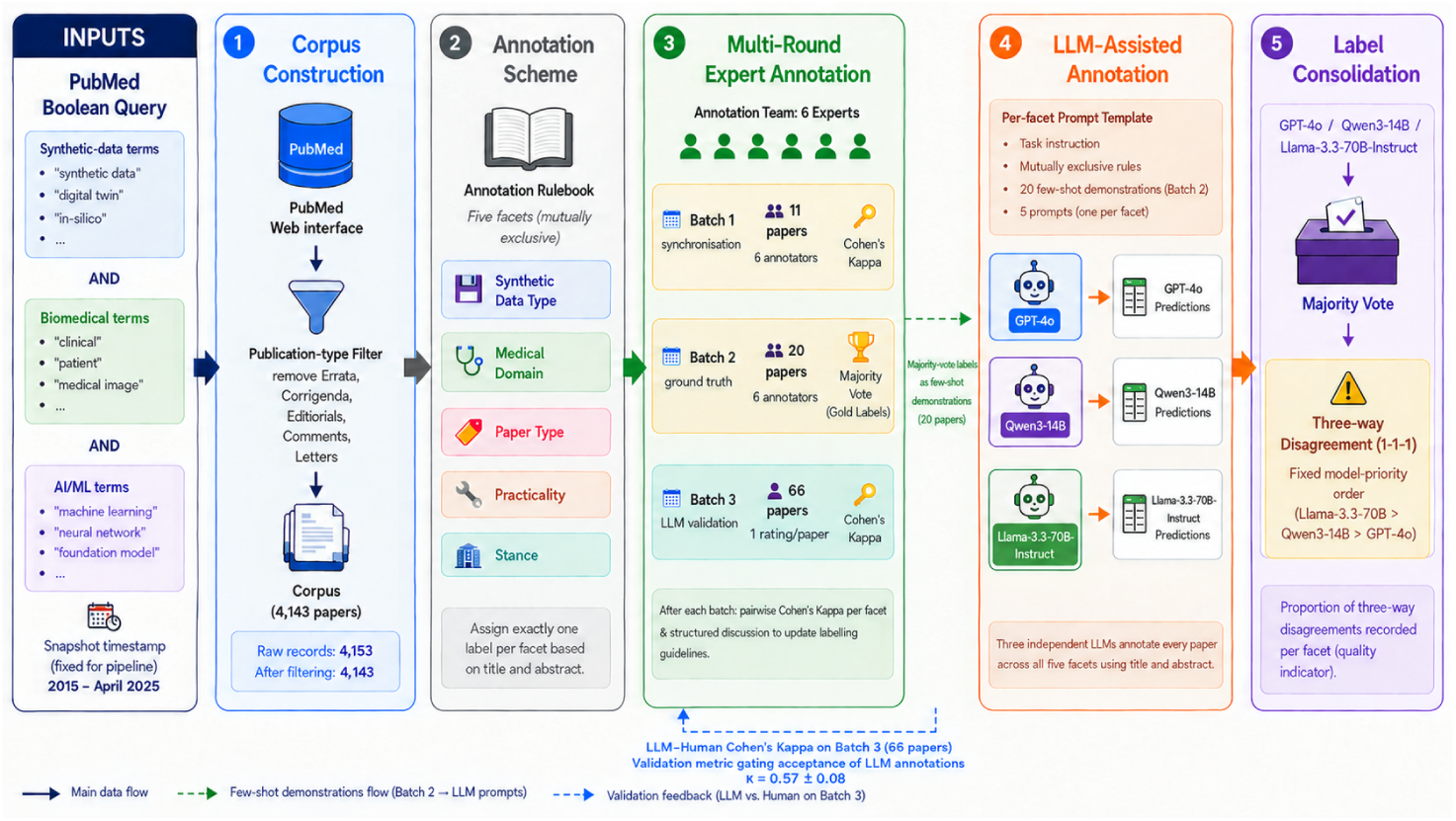
End-to-end annotation pipeline for the systematic mapping study. The pipeline proceeds through five sequential stages. (1) Corpus Construction: PubMed was queried in April 2025 using a structured Boolean query combining synthetic-data, biomedical-context, and AI-methodology term clusters; after automated exclusion of ten non-article entries, the corpus comprised 4,143 records (2015–April 2025). (2) Annotation Scheme: Each paper was assigned exactly one label per facet based on title and abstract. (3) Multi-Round Expert Annotation: Six independent experts annotated papers in three sequential batches; Cohen *κ* inter-rater agreement was computed pairwise after each batch and used to refine annotation guidelines. (4) LLM-Assisted Annotation: Three LLMs independently annotated all papers across all five facets using per-facet prompts with 20 few-shot expert-annotated demonstrations drawn from Batch 2; LLM–human agreement on Batch 3 served as validation metric. (5) Label Consolidation: Final labels were determined by majority vote across the three LLMs; three-way disagreements were resolved by a fixed model-priority order. Dashed arrows indicate the flow of Batch 2 majority-vote labels as few-shot demonstrations into the LLM prompts (green) and LLM–human inter-rater agreement (Cohen *κ*) feedback from Batch 3 (blue).

## The landscape of AI-based synthetic data in biomedicine

The corpus of 4,143 papers spanning 2015 to 2025 documents a field in sustained growth (*cf.* Figure 2A). Classification of papers according to the different facets is provided in Figure 3A-E. The *stance* facet distribution reflects a strongly productive research culture: 77.8% of papers (*N* = 3,225) are classified as strongly supportive of AI-based synthetic data approaches, while 8.6% express some degree of nuance or qualification (somewhat supportive, *N* = 356), and fewer than 1% (*N* = 33) adopt a critical stance. Notably, the number of critical papers rose from a handful before 2021 to several dozen after, representing a community beginning to interrogate methodological validity, evaluation standards, and generalization claims, a pattern consistent with field maturation in other applied AI domains.

**Fig. 2:**
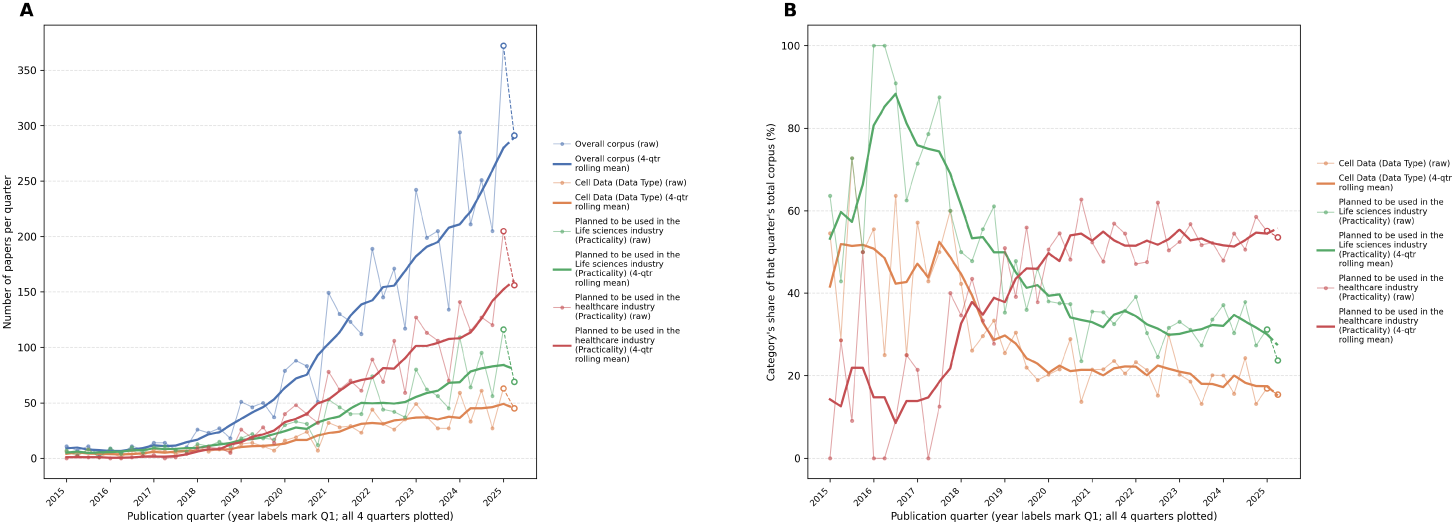
A: raw quarterly paper counts for the overall corpus and the three categories with the largest, FDR significant change in share of the corpus over time (Poisson interaction test, top three by effect size out of 18 candidates tested). B: the same three categories’ share of the overall corpus per quarter. Thick lines are a yearly (four-quarter) rolling mean; the final quarter (2025 Q2, open circles) reflects a single observed month and is shown as extrapolated.

**Fig. 3:**
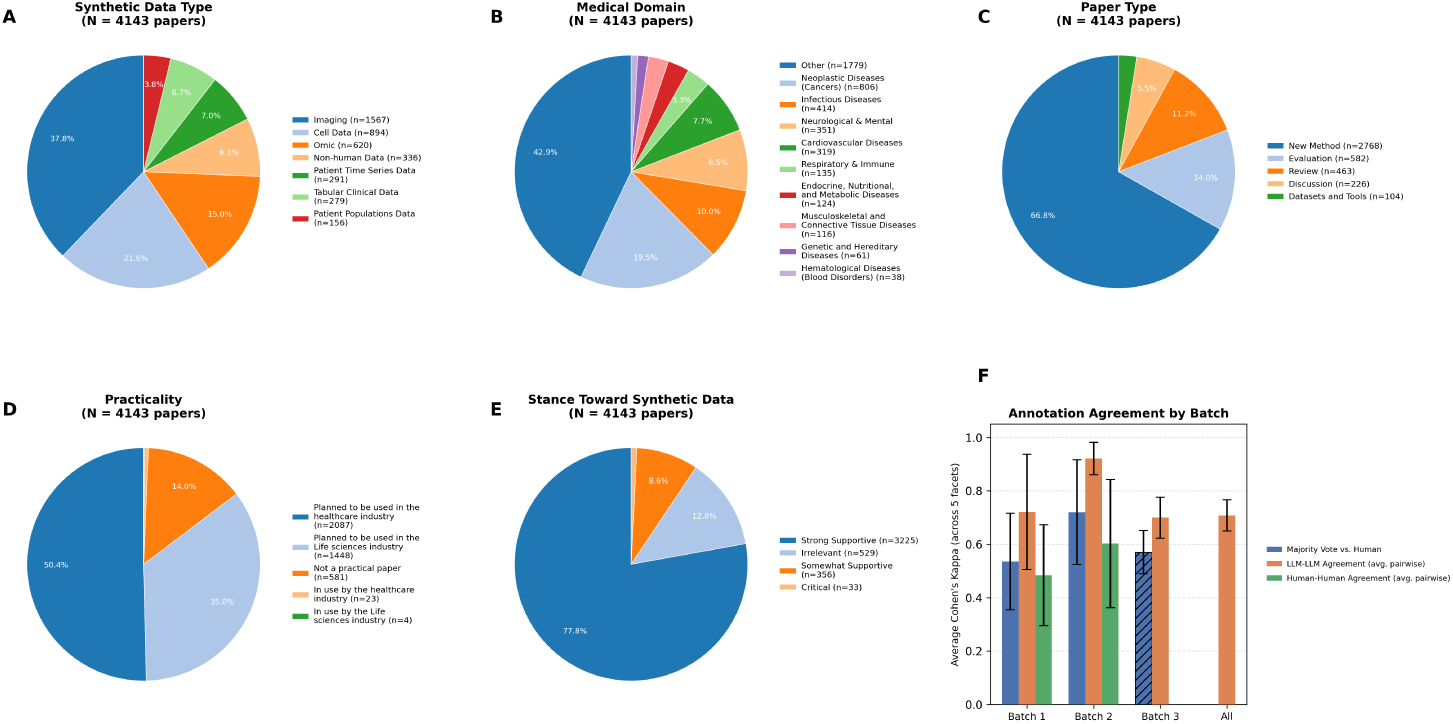
Facet distributions for the full corpus (*N* = 4,143 papers, majority vote labels across three LLM annotators). A through E: one pie chart per facet. F: interrater agreement across annotation batches, averaged over all five facets. Bars show Majority Vote *vs.* Human agreement, average pairwise agreement among the three LLM annotators, and average pairwise agreement among six human labelers (Batch 1 and 2 only). Error bars are one standard deviation across raters, except Batch 3’s Majority Vote *vs.* Human bar (hatched), a paper level bootstrap standard deviation, since Batch 3 has no second human rater to compare against.

Figure 2B focuses on the three categories with the most notable different temporal behavior than the overall corpus. It tracks each of the three categories’ share of all papers published in the same quarter, the same ratio the interaction test evaluates directly. The clearest pattern is a reordering between the two *practicality* categories.

Work aimed at the life sciences industry led early, reaching 90 *−* 100% of the quarter’s corpus in 2016 before easing to roughly 47 *−* 61% through 2018 (interaction coefficient *−*0.025, *q <* 0.001). Work aimed at the healthcare industry shows close to the mirror image: negligible before 2017, it first overtakes life-sciences-industry work in 2019 Q1 (51.0% versus 35.3%) and settles at roughly 47 *−* 58% of the corpus from 2021 onward (interaction coefficient +0.021, *q <* 0.001). Read together, these trends are consistent with life sciences applications, where data is typically less regulated and easier to obtain than clinical data, serving as an early proving ground for biomedical synthetic-data methods, with attention shifting toward the more heavily regulated healthcare setting as the underlying techniques matured. Cell Data (*Data Type*) follows a similar declining trajectory (54.5% of the corpus in 2015 Q1 to roughly 15*−*20% by 2024-2025; coefficient *−*0.027, *q <* 0.001), consistent with the corpus’s broader diversification away from its earliest, narrower focus.

Medical imaging dominates the *data-type* distribution (*N* = 1,567; 37.8%), consistent with the early and sustained adoption of GAN- and diffusion-based approaches for synthetic image generation in radiology, pathology, and medical photography. Patient-population data form the smallest corpus segment (*N* = 156; 3.8%) but carry the highest mean normalized citation rate of any data modality (1.04 citations/month), and non-human data (*N* = 336; 8.1%) show the widest dispersion of citations of any modality (*SD* = 2.96), reflecting a small number of very highly cited molecular resources rather than a uniformly high-impact segment.

New methodological papers constitute the largest *paper-type* category (*N* = 2,768; 66.8%), highlighting the strong emphasis on method development in this rapidly evolving field. For comparison, a similar analysis of explainable AI in healthcare and life sciences, using the same classification scheme but covering publications only through 2020, found that new-method papers represented the second-largest paper type and accounted for 16.8% of publications [26]. Although the two analyses do not cover the exact same time periods, this marked contrast underscores the exceptional level of methodological activity in synthetic biomedical data research, consistent with the rapid recent progress in generative AI and its broad and growing adoption across biomedical domains.

Citation analysis reveals a more nuanced picture that complicates the apparent imaging dominance. Among the ten most-cited papers in the corpus by normalized citation rate (Table 5), six are reviews or surveys, reflecting the well-known citation advantage of synthesis works: two focus on medical imaging [28, 29], one on drug discovery [30], and three cover broad methodological topics [8, 9, 31]. Of the four remaining non-review papers, three concern molecular and pharmaceutical applications (the CARD antibiotic resistance database [32], the ProTox toxicity prediction platform [33], and an AI-guided robotic platform for organic compound synthesis [34]), while only one addresses medical imaging [35]. Despite imaging’s dominance by publication volume, the highest-impact non-review contributions by citation velocity come disproportionately from the molecular and pharmaceutical domain, consistent with the strong real-world utility of these tools in drug discovery and life sciences workflows. The temporal evolution of research vocabulary, visualized in Figure 4, confirms the broader transition documented above. Traditional simulation- and image-processing-centered terminology is more characteristic of earlier publications, whereas generative AI and digital-twin terminology is concentrated in the post-2021 period. This pattern is consistent with the broader methodological shift in AI associated with the rise of transformer- and diffusion-based models.

**Fig. 4:**
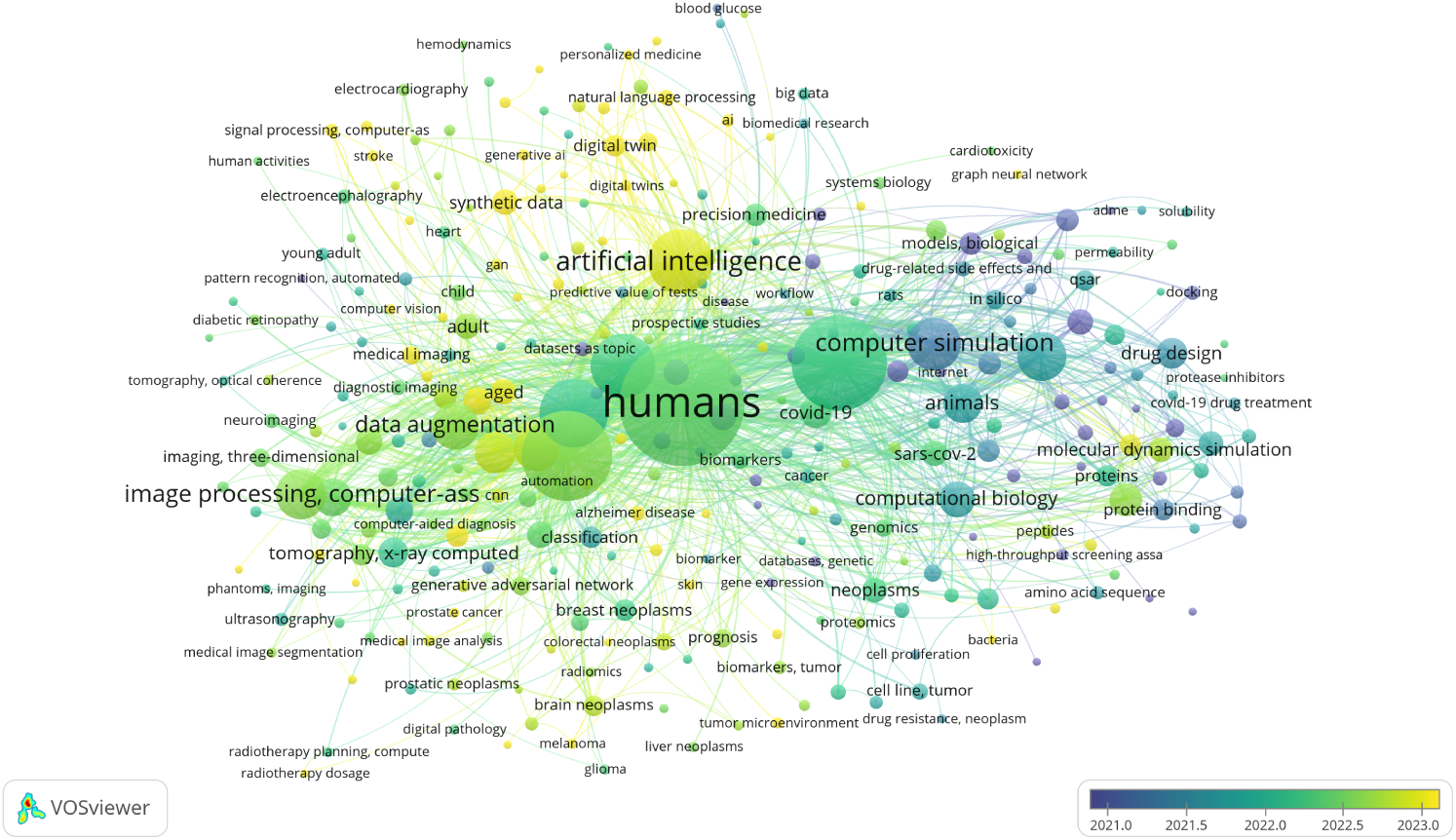
Term co-occurrence map created with VOSViewer [27]. Nodes represent the most frequent terms in titles and abstracts; node size reflects term frequency and link width reflects co-occurrence frequency between term pairs. Node color encodes the average publication year of articles containing that term, revealing temporal shifts in research focus: traditional methodological terms (*e.g.*, computer simulation, image processing) appear in earlier-dated articles on average, while terms such as Artificial Intelligence, Synthetic Data, and Digital Twin are concentrated in more recent publications (averaging around 2023). The displayed time range (2021–2023) reflects averaging over a corpus spanning 2015–2025, with the increased volume of recent publications skewing average years toward the later end of the window.

## Discussion and future directions

In this paper we analyzed ten years of systematic publication data, revealing a field that is large, fast-growing, and almost uniformly enthusiastic about its own methods. The most striking quantitative findings are: a *stance* distribution in which 86.4% of papers are supportive or somewhat supportive (77.8% and 8.6%, respectively) and fewer than 1% critical, with the critical minority emerging only from 2021 onward; a structural tension in terms of *data type* between imaging’s dominance by publication volume and the molecular and pharmaceutical domain’s dominance by citation impact among non-review papers; and a profound *practicality* gap: of 4,143 papers retrieved, only 27 carry an *In use* label, and a close reading of those papers reveals that even this small number overstates genuine deployment across most of the field, as several describe validation studies or proof-of-concept trials rather than routinely operational systems. A notable exception is drug discovery [36], where AI-based synthetic data tools such as *in silico* ADMET modeling and molecular synthesis platforms are already in routine industrial use, as reflected by several of the top-cited life sciences papers in our corpus [34, 37]. These patterns describe a field that is methodologically productive but translationally immature, and whose literature captures the early stages of the research pipeline far better than the later ones.

A related set of risks concerns the long-term epistemic consequences of the synthetic data paradigm itself. In natural language and vision, recursive training on model-generated data has raised concerns about AI autophagy: as synthetic datasets accumulate in public repositories and benchmarks, downstream models trained on mixtures of real and synthetic data may inherit and amplify distributional distortions, reducing diversity and degrading performance in ways that are difficult to detect from within the training pipeline [38, 39]. In biomedicine, where ground truth is expensive to acquire and validation against real patient outcomes is constrained by privacy and access, this risk is compounded. The growing prevalence of synthetic data in public biomedical datasets creates conditions in which data contamination and bias amplification could propagate silently. More broadly, the dominance of AI-generated surrogates for data, particularly in domains where they substitute for measurement rather than augment it, may subtly narrow the hypothesis space that researchers explore, fostering what has been described as illusions of understanding: predictive capability mistaken for mechanistic insight [40]. One partial response, again best understood as complementary rather than substitutive, is to fold mechanistic constraints directly back into the discovery loop, as in symbolic-regression frameworks that combine data with axiomatic background theory [41, 42]; the limits of such approaches at biomedical scale remain open, but they offer a route to keeping data-driven models accountable to known physics and chemistry. Our corpus finding that the critical literature remains below 1% of total output, and that it only gained traction from 2021, suggests that the community is only beginning to develop the evaluative infrastructure needed to interrogate these risks.

Several limitations bear on the interpretation of these findings. First, our corpus is restricted to PubMed-indexed records in English. Major venues for AI methodology, including arXiv, bioRxiv, medRxiv, NeurIPS, ICML, ICLR, MICCAI, and IEEE Transactions on Medical Imaging, are under-indexed in PubMed, and our analysis is therefore best understood as a characterization of the biomedically situated, peer-reviewed segment of the field rather than its full methodological frontier. Second, annotation reliability was moderate and varied across facets. Human inter-rater agreement improved following refinement of the annotation guidelines, *κ* = 0.60 *±* 0.24, and agreement between human annotations and the LLM majority-vote labels in the held-out Batch 3 test set was *κ* = 0.57 *±* 0.08. The reported corpus-level percentages should therefore be interpreted as broad field-level estimates rather than precise measurements of individual papers. Third, the 2025 portion of the corpus reflects records indexed through 30 April 2025 and is therefore an incomplete year, which we have flagged on the relevant time-resolved figures to prevent misreading of trends. Fourth, and conceptually most consequential (and a point on which the field will likely continue to disagree productively), the boundary of what constitutes “synthetic data” is itself unsettled. Our typology subsumes outputs as diverse as GAN- or diffusion-generated medical images, which are samples from a learned approximation of an empirical distribution; digital twins, which are mechanistic models conditioned on individual or population data; and predictions from structure-prediction systems such as AlphaFold, which are regressions to a physical observable conditioned on evolutionary and structural priors. These objects have different epistemic statuses. We have chosen to treat them as a single corpus for the purposes of describing the literature while distinguishing them in our discussion of impact, but a reader who applies a stricter definition of “synthetic” would arrive at different headline counts.

The above observations point to five concrete priorities for the next decade. First, the research community should rebalance what its evaluation studies evaluate. Evaluation is not scarce in absolute terms: it accounts for 14.1% (*N* = 582) of the corpus, but it is heavily concentrated on the efficacy of image augmentation, with the most-cited evaluation papers comparing augmentation techniques and architectures on imaging benchmarks. Systematic head-to-head comparisons of synthetic against real data on clinical endpoints, and on non-imaging modalities, remain comparatively rare; without them, deployment claims will remain difficult to adjudicate. Second, journal editors and funders should encourage longitudinal follow-up reports on high-citation tools (over cross-sectional citation analyses at a single point in time), analogous to the Bayer ADMET retrospective, that document post-publication adoption and real-world performance. Third, non-imaging data modalities, particularly tabular clinical data and omics, warrant targeted investment: the citation analysis suggests that high-impact contributions in these areas are achievable, but the volume of work remains disproportionately low relative to the clinical importance of the underlying *data types*. Fourth, geographic diversity in authorship and in the patient populations represented in training data must be treated as a quality criterion, not an afterthought, if synthetic data is to serve as a foundation for global healthcare AI.

A fifth and increasingly pressing priority is the development of synthetic-data-specific reporting standards. The biomedical AI community has converged on a set of reporting checklists for adjacent activities, including TRIPOD-AI [43], MINIMAR [44], FUTURE-AI [45], DECIDE-AI [46], SPIRIT-AI [47], and CONSORT-AI [48], none of which was designed for synthetic data specifically. Key elements central to the trustworthiness of synthetic data remain uncovered: the identity and version of the generative model, the provenance of its training data, the fidelity, utility, and privacy trade-offs characterizing the released sample, downstream task performance on a held-out real cohort, and the residual distributional shift between real and synthetic data. A focused community effort to extend existing checklists, or to draft a SPIRIT-AI-style standard for synthetic-data studies, would directly address the evaluation imbalance our corpus reveals. We are mindful of reporting-checklist fatigue in the field, but the gap here is specific enough and the downstream consequences large enough that a focused extension rather than a standalone framework seems warranted.

It is worth pausing on a conceptual distinction that the surveyed literature largely elides. AI-based synthetic data generation, as practiced in essentially all of the 4,143 records examined here, learns to approximate an empirical distribution from training samples and draws new observations from that learned approximation. A different strategy would be to derive synthetic observations from physical laws themselves (for instance, electronic-structure calculations on a target molecule, or open-system master-equation simulations of a biomolecular process), in which case the resulting datum is closer to a regression to a physical observable than to a draw from a fitted density. The validation strategy, and the conditions under which trust is warranted, differ in each case; this distinction strikes us as worth a careful taxonomy in future analyses. A related and under-appreciated caveat applies to physics-based generation more broadly: identifiability of an underlying generator from simulated or observed trajectories depends sharply on the ergodic properties of the dynamics, so a single simulated trajectory does not necessarily pin down the system the way one might assume [49].

An emerging direction worth highlighting is the use of quantum simulation as a source of physics-grounded synthetic observations, potentially in combination with generative AI. In this hybrid paradigm, portions of a molecular system where quantum effects are physically significant, for instance electronic structure at a catalytic active site, could be handled by quantum hardware, while generative models on classical GPU and CPU infrastructure handle the surrounding conformational or scaffold-level variation. This positions quantum simulation not as a replacement for generative synthetic data, but as a physics-informed upstream component that injects quantum mechanical signal into a broader generative pipeline. Current hardware faces well-recognized challenges, including barren-plateau effects that constrain trainability at scale, and biomedically relevant system sizes remain beyond near-term reach. Nevertheless, the conceptual case for this hybrid architecture is independent of current device limitations, and the trajectory of hardware development suggests it would benefit from keeping these two questions distinct.

The decade documented here represents a foundational period: the methods were established, the applications were mapped, and the scale of interest was confirmed. The next decade will be defined by whether the field can close the gap between what synthetic data promises and what it demonstrably delivers in the clinic, the laboratory, and the population.

## Methods: Systematic review methodology

### Literature search and corpus construction

For this analysis, we adopted a broad operational definition of AI-based synthetic biomedical data, encompassing biomedical observations generated or transformed using machine learning or generative AI methods, including data-driven generation and data augmentation. We conducted a selection process documented following PRISMA 2020 reporting principles [50], following an approach established in prior bibliometric analyses of biomedical AI [26, 51, 52]. The full pipeline is illustrated in Figure 1; Panel 1 (Corpus Construction) details the query structure and filtering steps.

Records were retrieved from a single bibliographic database, PubMed, through its advanced search web interface (https://pubmed.ncbi.nlm.nih.gov/advanced/). No other database, register, preprint server or grey-literature source was searched, and no reference-list or citation chasing was performed. The search was executed in April 2025, and every record analysed here derives from that single retrieval. The query required the co-occurrence of terms drawn from three clusters combined with AND, together with a disjunctive arm capturing virtual-patient work:

- *Synthetic-data and simulation terms*: “synthetic data” OR “digital twin” OR “insilico” OR “data augmentation”.
- *Biomedical context terms*: “medical image” OR “clinical” OR “patient” OR “biomedical” OR “drug” OR “healthcare” OR “pathology” OR “medical”.
- *AI and machine-learning methodology terms* : “generative AI” OR “AI” OR “machine learning” OR “supervised” OR “neural network” OR “generative adversarial” OR “foundation model” OR “large language model” OR “transfer learning”.

The combination applied was (synthetic-data term AND biomedical term AND AI/ML term) OR (“virtual patient” AND AI/ML term). Two limits were set within the interface: publication date from 2015/01/01 to 2025/04/30, inclusive, and language English. The query returned 4,153 records, which were exported in XML and parsed to extract the PubMed identifier, title, abstract, author keywords, MeSH terms, journal, publication year and DOI. Step-by-step retrieval instructions, including the term clusters above, are given in Protocol A [53]. The search strategy was not independently appraised using a formal instrument such as PRESS.

PubMed was selected as the search database because the objective of this review was to examine the use of synthetic data in biomedical science. While synthetic data methods are actively developed in computer science and machine learning research, such work often focuses on methodological innovation, algorithmic performance, or general-purpose benchmarks, and is not necessarily situated in biomedical research contexts. Our focus was, instead, on biomedical uptake and use: the types of biomedical data being synthesized, the purposes for which synthetic data is applied, the biomedical tasks or research settings in which it is used, and how its utility, validity, privacy, and limitations are evaluated in relation to biomedical applications. PubMed was therefore chosen because it indexes literature in medicine, biology, public health, and related biomedical domains, making it well aligned with the scope of this study.

### Eligibility criteria and study selection

Eligibility was determined by two deterministic operations: the Boolean query with its date and language limits, described above, and a filter on PubMed publication type. A record was eligible if it was indexed in PubMed, published between 1 January 2015 and 30 April 2025 inclusive, written in English, matched the query, and was a primary research article or a review. A record was excluded if its PubMed publication type identified it as an erratum, corrigendum, editorial, comment or letter, on the grounds that these are not primary research or review contributions.

No further screening was applied to titles and abstracts, and no full texts were retrieved: the query and the publication-type filter jointly constitute the selection procedure. Selection therefore involved no subjective judgement, and no duplicate screening, adjudication of disagreements or inter-rater statistics arise at this stage. Because a single database was searched, no deduplication step was required; the retrieved set was verified to contain no duplicate DOIs. Relevance to the research question was likewise not applied as a pre-screening filter. It was instead captured during annotation through the *stance* facet, whose *Irrelevant* category denotes a paper that takes no position toward the use of synthetic data; all records, including those so labelled, were retained in the corpus and are included in every reported distribution.

Of the 4,153 records returned by the query, ten were removed by the publication-type filter — errata or corrigenda (*n* = 3), editorials (*n* = 3), and short commentaries, letters and other non-article items (*n* = 4) — leaving a final corpus of 4,143 records published between 2015 and April 2025, inclusive, in English. No records were excluded at any subsequent stage. The flow of records is shown in Figure 5, and the corpus construction steps are summarized in Panel 1 of Figure 1. The relatively small number of records excluded reflects the high precision of the search strategy: because eligibility is carried almost entirely by the query itself, precision at retrieval substitutes for the screening effort that a broader query would have required.

**Fig. 5:**
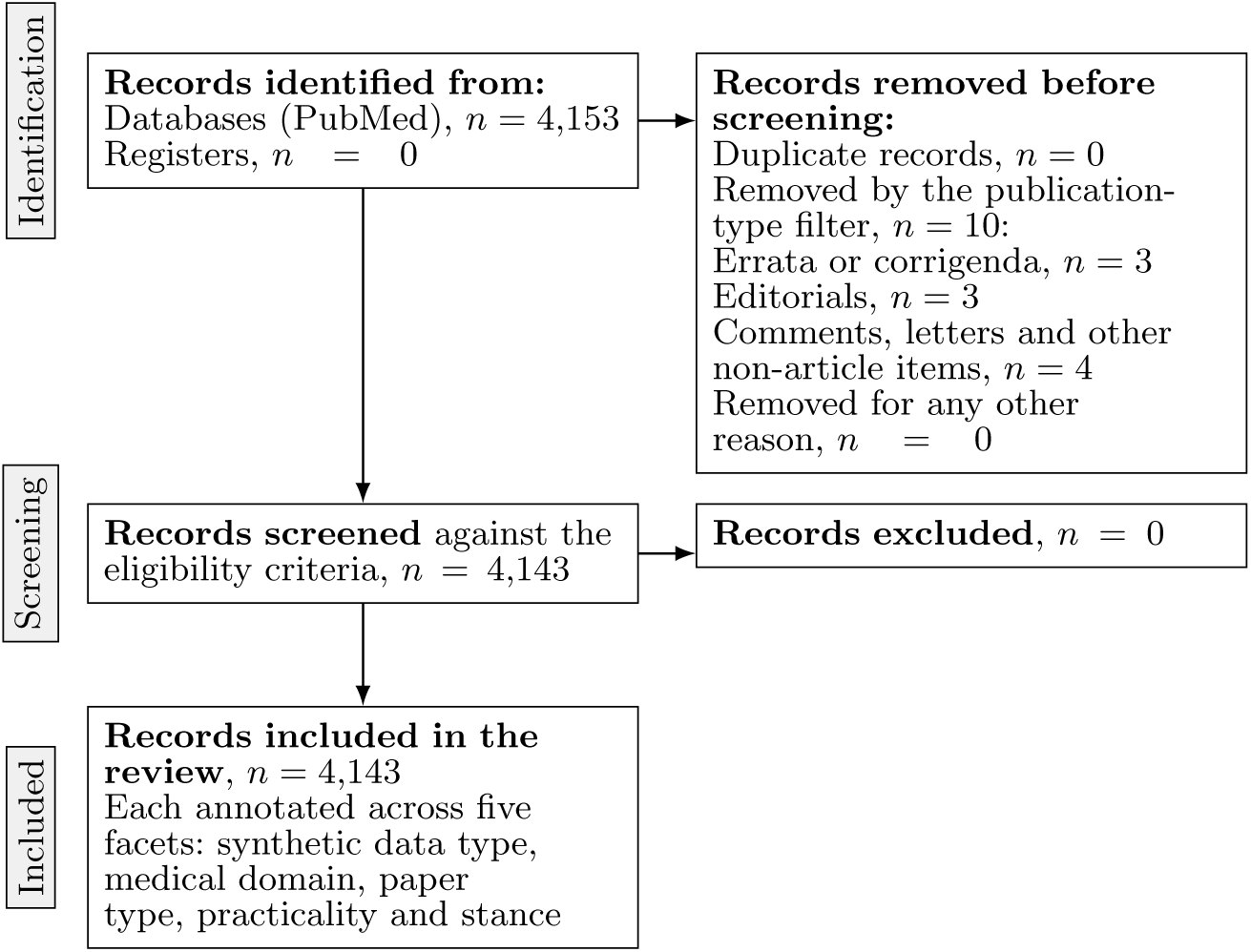
PRISMA 2020 flow diagram for corpus construction. Eligibility was determined by the Boolean query with its date and language limits together with a filter on PubMed publication type. No relevance screening was applied to titles and abstracts, which is why no records are excluded at the screening stage. No separate full-text eligibility stage is shown because none was performed: all data items were extracted from titles and abstracts. Papers taking no position toward the use of synthetic data were labelled *Irrelevant* within the *stance* facet during annotation and were retained in the corpus.

### Annotation, validation, and citation analysis

The complete search, screening, annotation, validation, and bibliometric procedures are available as step-by-step open protocols [53, 55] and illustrated across Panels 2–5 of Figure 1. Each paper was classified from its title and abstract across five facets: paper type, medical domain, synthetic data type, practicality, and stance toward synthetic data. Facet definitions and decision rules are summarized in Table 1 and detailed in the Supplementary Material.

**Table 1:** Summary of annotation facets. Full definitions and category lists are provided in Table 3.

| Facet | Definition | Categories |
| --- | --- | --- |
| 1. Paper Type | Type of scientific contribution, adapted from [26] | Review; Conceptual; Methods; Application; Data/tools |
| 2. Medical Domain | Disease area based on ICD-11 [54] | ICD-11 disease categories group into 10 groups |
| 3. Synthetic Data Type | Type of synthetic data generated or analyzed | Imaging; Omics; Clinical; Time series; Population; Cell; Non-human |
| 4. Practicality | Degree of real-world use or deployment | Not practical; Planned (in healthcare or life sciences); In use (in healthcare or life sciences) |
| 5. Stance | Position toward synthetic data usage | Supportive (strong or somewhat); Critical; Irrelevant |

Data extraction was performed against a fixed extraction sheet: each record was assigned exactly one value per facet, drawn from a closed vocabulary, on the basis of its title and abstract alone. Table 2 lists the extracted fields, their permitted values, and the provenance fields recorded alongside each label so that any value can be traced back to the annotators that produced it. The completed extraction sheet for all 4,143 records is deposited alongside the corpus (see Data availability).

**Table 2:** Data extraction sheet. One row per record; exactly one value is recorded per facet. Provenance fields are recorded for every record-facet pair so that each consolidated label can be traced to the individual annotators and to the rule that resolved it.

| Extracted field | Permitted values or format |
| --- | --- |
| *Bibliographic fields, taken from the PubMed XML export* |  |
| Record identifiers | PubMed identifier; DOI |
| Bibliographic metadata | Title; abstract; author keywords; MeSH terms; journal; publication year |
| Publication quarter | Year and quarter, parsed from the reference string; empty for the 107 records without a parseable month |
| *Facet values, extracted from title and abstract* |  |
| Synthetic data type | Imaging; Omic (genomics, proteomics, metabolomics); Tabular clinical data; Patient time-series data; Patient population data; Cell data; Non-human data |
| Medical domain | Neoplastic diseases (cancers); Infectious diseases; Genetic and hereditary diseases; Cardiovascular diseases; Neurological and mental disorders; Endocrine, nutritional and metabolic diseases; Musculoskeletal and connective tissue diseases; Hematological diseases; Respiratory and immune diseases; Other |
| Paper type | Review or meta-analysis; Conceptual discussion; Novel methods; Evaluation or application of methods; Datasets or tools |
| Practicality | Not practical; Planned for the healthcare industry; Planned for the life sciences industry; In use by the healthcare industry; In use by the life sciences industry |
| Stance | Strongly supportive; Somewhat supportive; Critical; Irrelevant |
| *Provenance fields, recorded per record-facet pair* |  |
| Per-annotator values | One value per LLM annotator, after normalisation to the canonical vocabulary of that facet |
| Resolution route | Unanimous; majority; tie-break; single source; no annotation |
| *Bibliometric fields, retrieved from OpenAlex* |  |
| Record linkage | OpenAlex work identifier; publication date |
| Citation measures | Cited-by count; retrieval date; months since publication; citations per month |

Data extraction was carried out in three sequential batches of records sampled at random and stratified by publication year. Six independent annotators with expertise in AI, data science, and medicine annotated 31 papers in Batches 1 and 2 with 11 and 20 papers, respectively. Following structured refinement of the annotation guidelines, mean inter-rater agreement improved from Cohen’s *κ* = 0.48 *±* 0.19 in Batch 1 to *κ* = 0.60 *±* 0.24 in Batch 2. For each record and facet, the batch consensus value was the modal value across annotators, with ties resolved by the most senior annotator in the relevant domain.

The 20 Batch 2 papers and their human expert majority-vote labels were used as few-shot examples for three LLM annotators: GPT-4o, Qwen3-14B, and Llama-3.3-70B-Instruct. Each model independently classified all 4,143 papers for each facet, with one API call per record, facet and model (4,143 *×* 5 *×* 3 = 62,145 calls in total).

Because the three models returned category names differing in spelling, capitalisation and abbreviation, each model’s output was first mapped onto a single canonical vocabulary per facet by case-insensitive matching after whitespace stripping; strings absent from the mapping were passed through and flagged rather than silently merged, and blank or missing responses were recorded explicitly. Final labels were then determined by majority vote across the three models. Where all three models disagreed, the label was resolved by a fixed model-priority order (Llama-3.3-70B *>* Qwen3-14B *>* GPT-4o); this affected 300 of the 20,715 paper-facet decisions (1.4%). The resolution route for every paper-facet pair — unanimous, majority, tie-break, single source or no annotation — was recorded alongside the final label as a quality indicator.

A third batch of 66 papers were independently annotated by the experts and used as a held-out test set, bringing the total number of human-annotated papers to 97. Agreement between the human annotations and the LLM majority-vote labels in Batch 3 was *κ* = 0.57 *±* 0.08, confirming adequate agreement and validating the design choice of using LLM-based annotators at scale in the analysis over the 4, 143 articles in the corpus (*cf.* below). Figure 3F presents mean Cohen’s *κ* for human–human, LLM–LLM, and human–LLM majority-vote comparisons. Full inter-rater statistics are reported in Supplementary materials.

Extraction across the full corpus was performed by the ensemble of LLM annotators; it was not independently duplicated by human annotators beyond the 97 human-annotated papers of Batches 1 to 3. The corpus-level percentages reported here should accordingly be read as field-level estimates rather than as accurate values for any individual record.

To identify which annotation categories grew or shrank at a rate distinguishable from the corpus overall, we restricted attention to candidate categories with at least 300 papers across the five facets (18 candidates). Each paper’s publication quarter was parsed from its Reference string; 107 papers (2.6% of the corpus) without a parseable month were excluded from this time-resolved analysis only and retained for all other analyses. For each candidate category, we fit a Poisson regression of quarterly paper counts on a centered quarter index, category membership (that category versus the rest of the corpus), and their interaction, over the 41 complete quarters from 2015 Q1 to 2025 Q1. The interaction coefficient captures the category’s excess quarterly growth rate relative to the rest of the corpus, independent of the corpus’s own overall growth. Raw p-values across the 18 candidates were corrected for multiple comparisons with the Benjamini-Hochberg false discovery rate procedure (*q <* 0.05); six categories were significant at this threshold, and the three with the largest absolute effect size are shown in Figure 2B. The final quarter (2025 Q2) reflects a single observed month rather than a full quarter; it was excluded from model fitting and is shown as an extrapolated, open-circle tail in Figure 2A for descriptive context only.

Bibliographic metadata were obtained by resolving each PubMed identifier to an OpenAlex record through the OpenAlex API; identifiers that did not resolve automatically were matched by manual title search. Citation counts were retrieved from the OpenAlex API on 6 November 2025, pinned to that single date, and normalized by the number of months since publication, computed as elapsed days divided by 30.44. The complete bibliometric pipeline, including citation-rate normalization and per-facet impact analyses, is described in Protocol B [55]. Additional methodological details are provided in the Supplementary Materials.

Five separate prompts were designed (one per annotation facet) each structured identically: a task instruction; the complete set of category options with definitions; and 20 few-shot demonstration examples drawn from the Batch 2 annotated papers, each comprising a title, abstract, assigned label, and brief reasoning. Prompts were applied in a per-facet, per-paper manner, and a separate API call was made for each facet, rather than attempting to classify all facets simultaneously. This design allowed independent quality control per dimension and simplified the output parsing. Three LLMs (GPT-4o, Qwen3-14B, Llama-3.3-70B-Instruct) were used as annotators. Final labels were determined by majority vote across the three models, with three-way disagreements resolved by the fixed model-priority order given above. Across the five facets, between 58.2% (stance) and 84.6% (medical domain) of papers received unanimous labels from all three models; the share of papers decided by a split 2-of-3 vote is itself a useful index of facet difficulty. The inter-rater annotation consistency across the 3 LLMs for each of the batches as well as the average agreement and standard deviation between human and LLM for batches 1, 2 and 3 are summarized in Figure 3F.

## Supplementary information

### Facet full list

Each paper was evaluated across five classification dimensions, or facets, based on its title and abstract. The classification scheme was adapted from prior biomedical AI systematic reviews [26] and extended to capture the specific characteristics of synthetic data research. The five facets, their definitions, and category sets are summarized in Table 3.

**Table 3:** Detailed annotation facets and classification scheme. Each paper was annotated across five dimensions. Categories are listed in full to ensure reproducibility of the labeling process and transparency of the classification criteria.

| Facet | Definition | N<sub>cat</sub> | Categories |
| --- | --- | --- | --- |
| **Paper Type** | Type of scientific contribution (adapted from prior biomedical AI reviews) | 5 | Review or meta-analysis; Conceptual discussion; Novel methods; Evaluation or application of methods; Datasets or tools |
| **Medical Domain** | Disease area, based on ICD-11 classification | 10 | Neoplastic diseases (cancers); Infectious diseases; Genetic and hereditary diseases; Cardiovascular diseases; Neurological and mental disorders; Endocrine, nutritional and metabolic diseases; Musculoskeletal and connective tissue diseases; Hematological diseases; Respiratory and immune diseases; Other |
| **Synthetic Data Type** | Type of synthetic data generated or analyzed | 7 | Imaging; Omic (genomics, proteomics, metabolomics); Tabular clinical data; Patient time-series data; Patient population data; Cell data (*e.g.*, single-cell RNA sequencing, microscopy); Non-human data |
| **Practicality** | Degree of real-world use or deployment | 5 | Not practical; Planned for the healthcare industry; Planned for the life sciences industry; In use by the healthcare industry; In use by the life sciences industry |
| **Stance** | Position toward the use of synthetic data | 4 | Strongly supportive; Somewhat supportive; Critical; Irrelevant |

*Note:* N<sub>cat</sub> = number of categories per facet. Full definitions and decision rules for each category are provided in Protocol A [53].

### Expert annotation and inter-rater reliability

Six independent reviewers with diverse expertise in AI, data science, and medicine manually annotated the papers in three sequential batches across five dimensions. The annotation workbench consisted of a shared spreadsheet in which reviewers recorded labels for each paper and dimension based solely on the title and abstract.

Batch 1 (*N* = 11 papers; all six annotators) served as an initial synchronization exercise. Inter-rater agreement was moderate, as measured using Cohen’s *κ* coefficient [56]. See Figure 6 for a summary of inter-rater agreement results. These findings prompted a structured discussion to refine and clarify the labeling guidelines. Key ambiguities resolved during this process included: (i) the inclusion of *in silico* modeling approaches within scope (aligned with the *in silico* medicine definition the use of computational models and simulations to represent biological processes, disease progression, or therapeutic interventions); (ii) the distinction between healthcare and life sciences industry practicality categories; and (iii) the boundary between data augmentation and synthetic data generation. The relevance-related discussion also led to a key decision: papers using *in silico* terminology were included if the approach contributed to disease prevention, diagnosis, prognosis, treatment, or management.

**Fig. 6:**
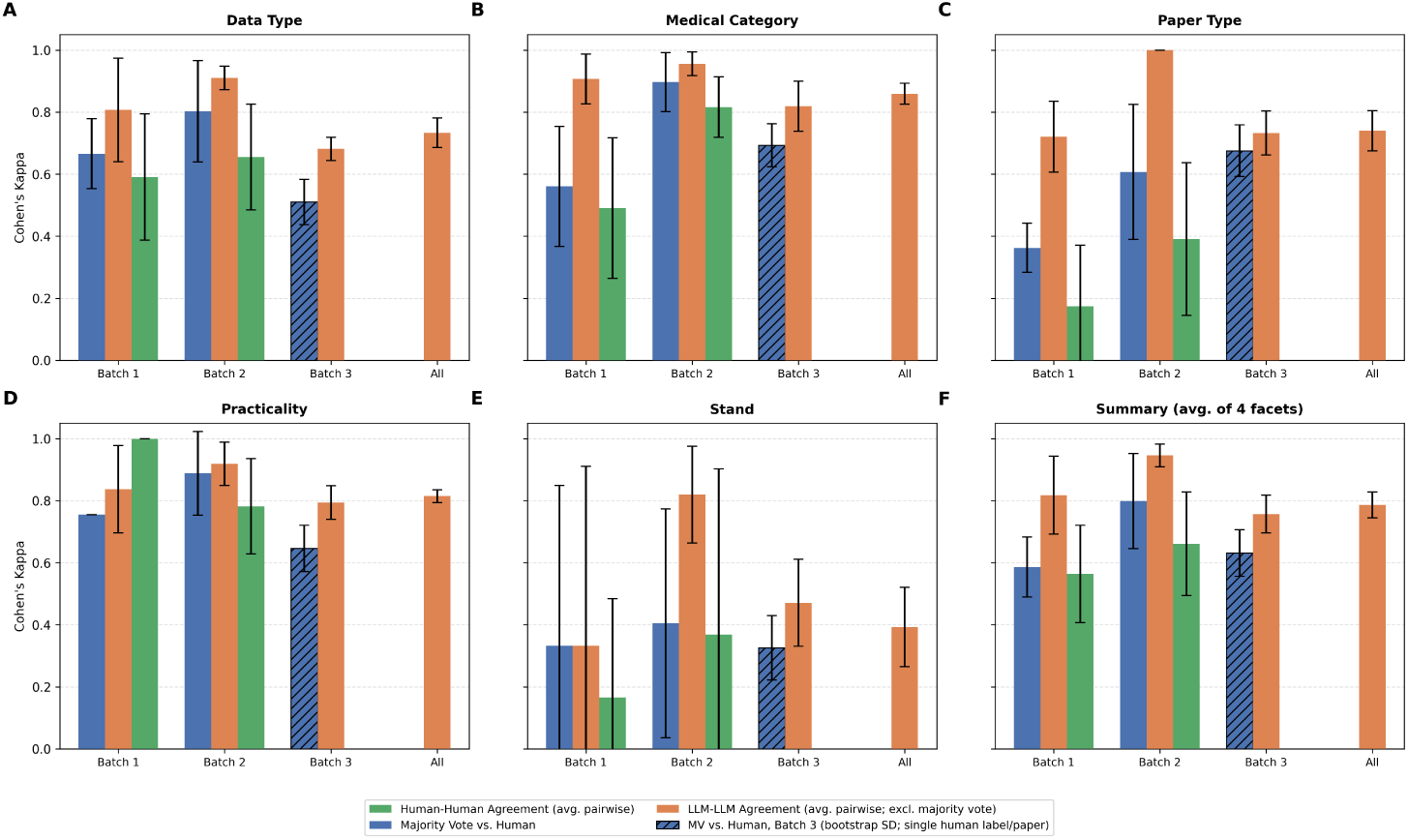
Cohen’s *κ* inter-rater agreement across annotation facets and batches. Panels A–E show pairwise Cohen’s *κ* for each of the five annotation facets: (A) Synthetic Data Type, (B) Medical Domain, (C) Paper Type, (D) Practicality, and (E) Stance. Panel F shows the average Cohen’s *κ* across all facets except Stance. Within each panel, agreement is shown for Batch 1 (synchronization, N = 11), Batch 2 (ground truth for few-shot demonstrations, N = 20), Batch 3 (LLM validation, N = 66), and the full annotated set (All). Four measures are displayed: Human–Human Agreement (average pairwise Cohen’s *κ* among the six human annotators; blue), LLM–LLM Agreement (average pairwise Cohen’s *κ* among the three LLM annotators; orange), Majority Vote *vs.* Human (Cohen’s *κ* between the LLM majority-vote label and individual human annotators; green), and MV *vs.* Human for Batch 3 (hatched dark bars). Error bars represent one standard deviation across rater pairs; for the Batch 3 Majority Vote *vs.* Human bar (hatched), a paper-level bootstrap standard deviation is used since Batch 3 has no second human rater to compare against.

Batch 2 (*N* = 20 papers; all six annotators) was annotated following guideline refinement. These 20 papers, together with their majority-vote labels among annotators across all five dimensions, were subsequently used as few-shot demonstration examples in the large language model (LLM) prompts. Human–human agreement computed on Batch 2 improved to Cohen’s *κ* = 0.60 *±* 0.24 across the five retained facets, representing a moderate to substantial level of agreement suitable for downstream use by the LLMs as ground truth .

### Inter-Rater Reliability and Annotation Refinement

Six independent reviewers (GEU, IFP, LZ, MMM, MRZ, NA) with diverse expertise (AI, data, medicine) manually labeled a subset of *∼*100 articles based on title and abstract. To evaluate consistency and reliability, we conducted an inter-rater agreement analysis using Cohen’s *κ* (Kappa) statistic [56]^1^ [^1^*sklearn* was used to compute Cohen’s *κ*. Other metrics (*e.g.*, Fleiss’s *κ*[57], Krippendorff’s *α* [58]) were considered but were revealed to have minimal impact. Cohen’s *κ* was chosen for its wide recognition, simplicity, and ease of interpretation (range: *−*1 to +1, where 1 indicates complete agreement, 0 chance-level agreement, and *−*1 complete disagreement). See [59] for details.]. Cohen’s *κ* measures agreement between independent evaluations of categorical variables while accounting for chance^2^ [^2^The sample size of 20 articles for inter-rater analysis was based on prior guidelines [60], which recommend 11–28 samples. As our goal was to assess relative improvement after guideline refinement, exact sample size was not critical.].

We aimed to map papers by AI technology type —pre-GenAI or GenAI; however, raters showed high variability, likely due to differences in AI expertise as well as the limited information available in an abstract. Labeling the type of AI technology is a challenging mission that has been addressed by different groups using different approaches, *e.g.* [52]). The multi-round annotation and inter-rater reliability procedure is captured in Protocol A [53]. The variabality per facet is captured in Figure 6.

After Batch 1, the human labelers discussed their labeling approach, reflected in the higher human to human *κ* in Batch 2 (0.60 *vs.* 0.48). Batch 2’s (human) majority vote labels were also used as few shot examples in the LLM prompts, and majority vote to human agreement is correspondingly highest in Batch 2 (0.72) compared to Batch 1 (0.54) and Batch 3 (0.57).

All three differences were tested with a paper-level bootstrap procedure and remained significant after Benjamini-Hochberg correction for multiple comparisons (*q <* 0.05; Table 4): majority vote to human *κ* was higher in Batch 2 than Batch 1 (Δ*κ* = 0.184, 95% CI [0.008, 0.345], *q* = 0.043) and than Batch 3 (Δ*κ* = 0.150, 95% CI [0.003, 0.268], *q* = 0.043), and human-to-human *κ* improved from Batch 1 to Batch 2 (Δ*κ* = 0.118, 95% CI [0.005, 0.229], *q* = 0.043), see Table 4.

**Table 4:**
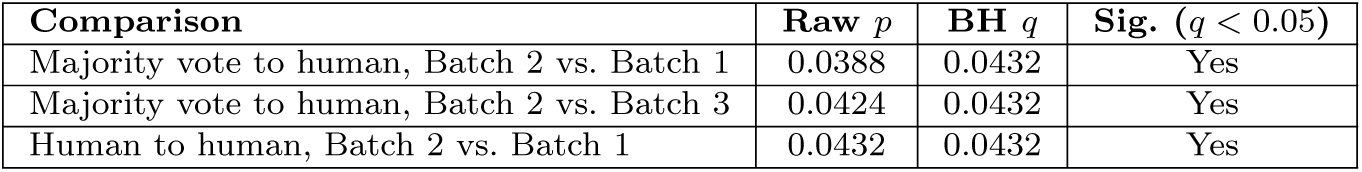
Paper level bootstrap tests for batch to batch differences in Cohen’s *κ*, with Benjamini-Hochberg FDR correction (*m* = 3). All three remain significant at *q <* 0.05; under the stricter Bonferroni correction, none would (adjusted *p >* 0.11 for all).

| Comparison | Raw $p$ | BH $q$ | Sig. ($q < 0.05$) |
| --- | --- | --- | --- |
| Majority vote to human, Batch 2 vs. Batch 1 | 0.0388 | 0.0432 | Yes |
| Majority vote to human, Batch 2 vs. Batch 3 | 0.0424 | 0.0432 | Yes |
| Human to human, Batch 2 vs. Batch 1 | 0.0432 | 0.0432 | Yes |

**Table 5:**
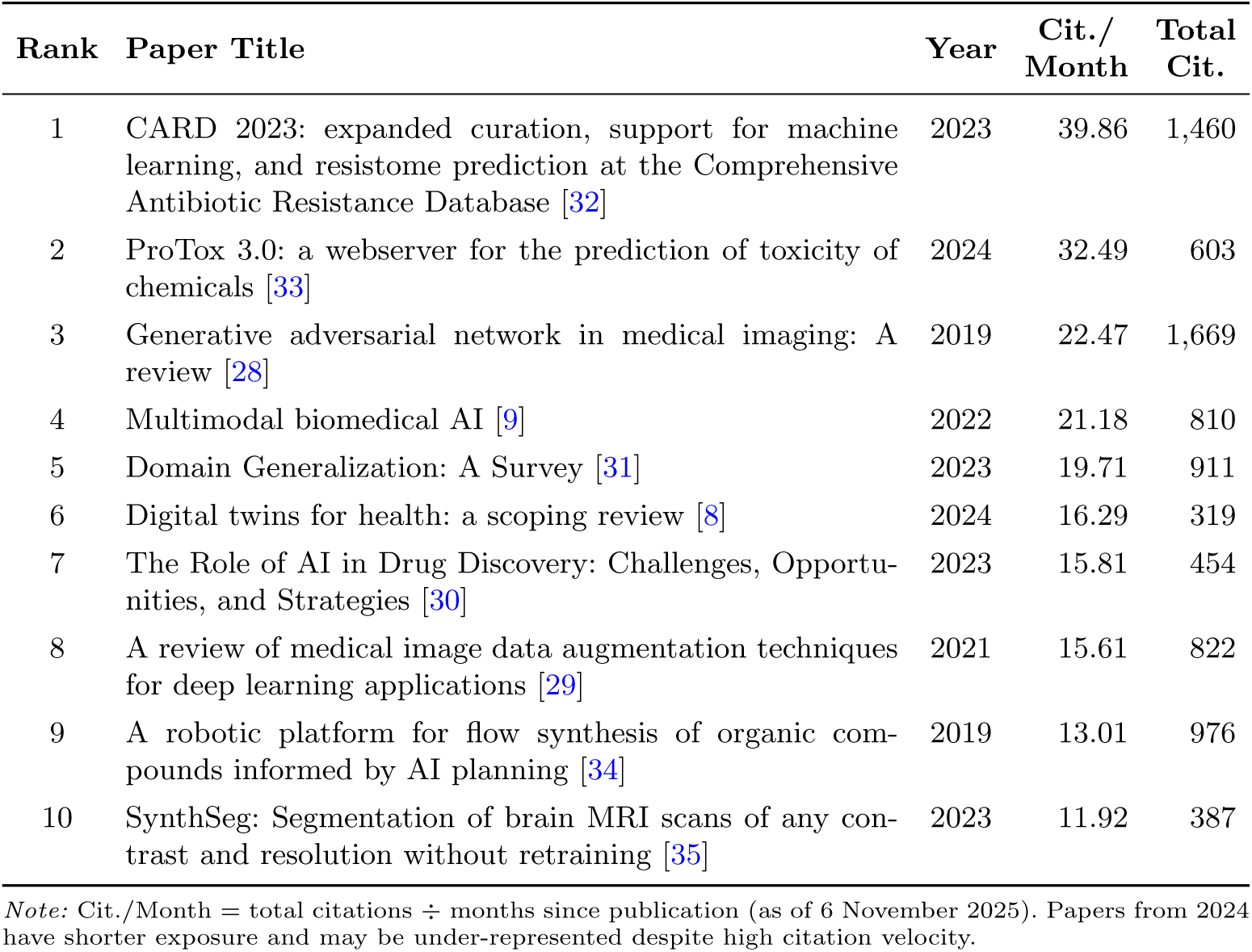
Top 10 papers in the corpus ranked by normalized citation rate (citations per month, computed as total citations divided by months elapsed since publication, as of 6 November 2025). Papers are ordered by descending normalized citation rate across all 4,143 papers retrieved from PubMed (2015–2025).

| Rank | Paper Title | Year | Cit./<br>Month | Total<br>Cit. |
| --- | --- | --- | --- | --- |
| 1 | CARD 2023: expanded curation, support for machine learning, and resistome prediction at the Comprehensive Antibiotic Resistance Database [32] | 2023 | 39.86 | 1,460 |
| 2 | ProTox 3.0: a webserver for the prediction of toxicity of chemicals [33] | 2024 | 32.49 | 603 |
| 3 | Generative adversarial network in medical imaging: A review [28] | 2019 | 22.47 | 1,669 |
| 4 | Multimodal biomedical AI [9] | 2022 | 21.18 | 810 |
| 5 | Domain Generalization: A Survey [31] | 2023 | 19.71 | 911 |
| 6 | Digital twins for health: a scoping review [8] | 2024 | 16.29 | 319 |
| 7 | The Role of AI in Drug Discovery: Challenges, Opportunities, and Strategies [30] | 2023 | 15.81 | 454 |
| 8 | A review of medical image data augmentation techniques for deep learning applications [29] | 2021 | 15.61 | 822 |
| 9 | A robotic platform for flow synthesis of organic compounds informed by AI planning [34] | 2019 | 13.01 | 976 |
| 10 | SynthSeg: Segmentation of brain MRI scans of any contrast and resolution without retraining [35] | 2023 | 11.92 | 387 |

*Note:* Cit./Month = total citations ÷ months since publication (as of 6 November 2025). Papers from 2024 have shorter exposure and may be under-represented despite high citation velocity.

### Top cited papers

Bibliographic metadata were retrieved for all papers in the corpus using the OpenAlex API, mapping PubMed identifiers to OpenAlex records. Citation counts were retrieved as of 6 November 2025. To account for differences in publication age, we computed a normalized citation rate defined as total citations divided by months elapsed since publication. For each facet and category, we computed summary statistics (mean, median, interquartile range). The ten papers with the highest normalized citation rate across the corpus were identified (Table 5), providing a qualitative face-validity check on the LLM-assigned labels. Citation measures were computed as specified in Protocol B [55].

## Data Availability

The corpus was derived from PubMed records retrieved in April
2025 (publicly available at https://pubmed.ncbi.nlm.nih.gov); bibliographic meta-data was obtained from OpenAlex (https://openalex.org) via its public API. The annotated corpus generated in this study, comprising the 4,143 records with their five-facet labels and the derived normalized citation-rate tables, has been deposited in Zenodo at https://doi.org/10.5281/zenodo.21720346. The expert-annotated batches used as ground truth and few-shot examples are provided in Zenodo at https://doi.org/10.5281/zenodo.21720346. Procedures for corpus construction, annotation, and bibliometric analysis are described in the associated protocols

## Acknowledgments

We acknowledge the BDVA (not-for-profit organisation boosting data and AI research, development and innovation for European competitiveness, societal wellbeing and sustainable progress), for facilitating discussions and collaborations that contributed to this work. We thank Eitan Farchi for helpful discussions. For transparency, as per ICMJE disclosure policy and as stated in the body of the article, this work includes LLM assistance in the annotation of the corpus of articles, following few-shot demonstrations from expert labelers and later validated *via* inter-rater agreement measures with a separate test set (Batch 3).

## Author contributions

The authors made the following contributions. **Navid Asgari**: Conceptualization; Methodology; Software; Formal analysis; Investigation; Data curation; Funding acquisition. **Iñaki Ferńandez Pérez**: Conceptualization; Methodology; Formal analysis; Investigation; Data curation; Writing - original draft; Writing - review & editing; Visualization. **Gorka Epelde**: Conceptualization; Methodology; Protocol development; Formal analysis; Investigation; Data curation; writing - original draft; Writing - review & editing; Visualization. **Linghan Zhang**: Conceptualization; Methodology; Formal analysis; Investigation; Data curation; Writing - original draft; Writing - review & editing; Visualization. **Lior Horesh**: Methodology; Visualization; Writing - review & editing; **Carl Saab**: Writing - review & editing; **Mordechai Muszkat**: Conceptualization; Data curation; Methodology; Writing - review & editing; **Michal Rosen-Zvi**: Conceptualization; Data curation; Methodology; Formal analysis; Investigation; Data curation; Writing - original draft; Writing - review & editing; Project administration.

## Data availability

The corpus was derived from PubMed records retrieved in April 2025 (publicly available at https://pubmed.ncbi.nlm.nih.gov); bibliographic metadata was obtained from OpenAlex (https://openalex.org) *via* its public API. The annotated corpus generated in this study, comprising the 4,143 records with their five-facet labels and the derived normalized citation-rate tables, has been deposited in Zenodo at https://doi.org/10.5281/zenodo.21720346. The expert-annotated batches used as ground truth and few-shot examples are provided in Zenodo at https://doi.org/10.5281/zenodo.21720346. Procedures for corpus construction, annotation, and bibliometric analysis are described in the associated protocols [53, 55].

## Code availability

Records were retrieved through the PubMed advanced search web interface, so no custom code was used for retrieval. All custom code used in this study —majority-vote label consolidation, inter-annotator agreement, temporal trend analysis, facet distributions, and normalized citation-rate computation— has been deposited in Zenodo with a persistent DOI at https://doi.org/10.5281/zenodo.21720346. The corresponding human-readable protocols are available at protocols.io [53, 55].

